# Identification of splicing defects caused by novel exonic mutations in *LEF1* in Sudanese B- chronic lymphocytic leukemia patients: computational approach

**DOI:** 10.1101/2022.07.18.22277759

**Authors:** Elkhazin Ali Abd Elmageed Eltayeb, Fatelrhman Mahdi Gameel, Abeer Babiker Idris, Enas dk. Dawoud, Hajir Sir Elkhatim Hamid Mukhtar, Abdelmarouf Hassan Mohieldein, Mehmet Burak Mutlu, Ismail A. Elrhman Mohmmed Ali, Alsadig Gassoum, Mohamed A. Hassan

## Abstract

**Background:** Lymphocyte enhancer factor-1 (LEF-1) is a member of the LEF-1/TCF family of transcription factors that are critically involved in canonical Wnt/β-catenin Signaling to regulate B Lymphocyte proliferation and survival. Alteration of *LEF1* expression and function leads to leukemogenesis as well as other several neoplasms.

**Aims:** to identify mutations in exons two and three of the *LEF1* among B-CLL Sudanese patients. Also, to functionally analyze the detected SNPs using different in silico tools.

**Materials and methods:** Immuno-phenotype for the detection of B cells CD5 and CD19 markers was performed on 128 B-CLL Sudanese patients by using a flow cytometry technique. DNA extraction, conventional PCR, and Sanger sequencing were applied to the LEF1 gene. Also, we performed a mutational analysis for identified SNPs using bioinformatics tools.

**Results:** A positive CD5 & CD19 expression was found in B-CLL patients. No mutation was observed in exon two. While four mutations were observed in exon three; two of them were not reported in previous studies. Interestingly, splicing analysis predicted that these mutations could lead to splicing defects in *LEF1* pre-mRNA due to their potential effects on splicing regulatory elements (i.e. ESE).

**Conclusion:** the two mutations Pro134Pro and Ile135Asn (novel mutation) were detected in all enrolled CLL patients and they could be used as diagnostic and/or prognostic markers for CLL. Therefore, further in vitro and in vivo functional studies with a large sample size are required to verify the splicing effect of the detected mutations in *LEF1* pre-mRNA.

## 1. Introduction

B cell chronic lymphocytic leukemia (B-CLL) is a hematological malignancy characterized by abnormal proliferation and accumulation of mature B lymphocytes in the blood, bone marrow, and lymphoid tissues. It is a conventional example of human malignancies that primarily involves defects in the inducement of programmed cell death. However, there’s a lot of cellular and molecular studies have discovered that the gene function of B-CLL cells is optimally systematic to avoid apoptosis; and the patients with B-CLL have an apparent prevalence of autoimmune phenomena (autoantibodies against self-antigens) (1, 2).

B-CLL is the most common type of leukemia in adults, comprising 25% of all cases diagnosed in North America and Europe and more than 20000 new cases estimated in the United State in 2018. The incidence of CLL increases with age as the median age of 55 years and is more common in males than females (male: female ratio 2:1). It varies geographically and is much less common in the far East (3, 4). Furthermore, evidence showed that the incidence of CLL also varies by race; for instance, in the United States Caucasians race has a higher incidence compared to African Americans or Asian Pacific Islanders (5). Along with that incidence of CLL is far less in Asian countries such as China and Japan, where it is estimated to occur at a frequency, approximately, 10 percent of that is seen in Western countries (6). However, the incidence of CLL cases in Africa is higher when compared to Asia (7). Genetic rather than environmental factors are the most likely explanation for these differences. A genetic effect on incidence was initially suggested by observational studies that have shown Japanese who settled in Hawaii do not have a higher incidence of CLL than native Japanese (8). Further support for a genetic effect was provided by a genotyping study in African Americans which demonstrated that this population had a lower frequency of single nucleotide polymorphisms associated with an increased incidence of CLL in other populations (9). There are no clearly discernible occupational or environmental risk factors that predispose to CLL (10). Although there is no available proof of genetic transmission of this disease, certain genetic polymorphisms may predispose patients to familial cancer including CLL, for example polymorphisms in Lymphocyte enhancer factor-1 (*LEF-1*) gene (11, 12).

*LEF-1* is a member of the *LEF*-1/TCF family of transcription factors which is critically involved in canonical Wnt/β-catenin Signaling to Regulate B Lymphocyte proliferation and survival (13-15). Alteration of LEF1 expression and function lead to leukemogenesis as well as other several neoplasms (14, 16-18). Gutierrez *et al*. identified that the transcription factor lymphoid enhancer binding factor-1 (LEF1) along with the Wnt pathway has a pathogenic role in CLL (19). This explains its overexpression in B CLL compared to normal B cells (20). Moreover, it is associated with poor prognosis in CLL and has been considered as an effective biomarker in such type of leukemia (21-23). Therefore, studying mutations in *LEF1*, which could lead to losing of protein features, splice-site changes and/or changes that might affect the amount of mRNA is of great significance. In Sudan, although there are many symptomatic and progressive B CLL cases, to our knowledge, no previous studies have addressed the mutations in *LEF1* and its relation with CLL. Accordingly, this study aimed to identify mutations in exons two and three of *LEF1* gene, the hotspot regions in leukemia (24), among B-CLL Sudanese patients. Also, functional analyses of the detected SNPs were performed using different in silico tools. The analyses predicted the effect of these mutations on LEF1 protein structure and function, splicing process and the pathogenicity of the mutations. In addition, homology modeling and 3D structure of LEF1 protein was predicted using raptorX and Chimera software.

## 2. Materials and Methods

### 2.1 Study setting and study population

This descriptive cross-sectional study was carried out in undertaken Radiation and Isotopes Centre of Khartoum (RICK) during the period of March 2014 to August 2017. One hundred and twenty-eight patients diagnosed as CLL were recruited from the (RICK). A structured questionnaire was used to obtain demographic & social information, and type of treatment from participants.

### 2.2 Sample collection

Five ml of the blood sample was taken from the capital vein in Ethylene-diamine-tetra-acetic acid (EDTA) (1.2mg/ml blood) anticoagulant.

### 2.3 Hematological analyses

A complete blood count test (CBC) for red blood cells, white blood cells Hemoglobin, Hematocrit, Platelets using SysmexKx 21N (Sysmex Corporation; Mundelein, llinois, Sysmex America, Inc) was done for all samples from patients.

### 2.4 Immunophenotypic analyses

20 µL of monoclonal florescent labeled antibodies was added into each labelled tube for flow cytometry analysis, then 100 µL of blood sample was added containing no more than 1 × 10 leukocytes / ml. (Counted by hematology analyzer - SYSMEX). Each tube was vortexed for 5 seconds and incubated at room temperature (18-25 OC) for 15 minutes. One ml of the “fix-and-lyse” mixture was added to the tube and vortexed immediately for three seconds, each tube was incubated at room temperature for at least 10 minutes and under the dark light. The tube was centrifuged at 150 × g for 5 minutes and the supernatant was discarded by aspiration, then 3 mL of phosphate buffer saline (PBS) was added to the remain of the test tube. All tubes were centrifuged at 150 × g for 5 minutes and the supernatant were discarded by aspiration. The pellets were re-suspended by addition of 0.5 to 1 mL of 0.1% formaldehyde. Then, all tubes were vortex for 5 seconds. Finally, flow cytometry was performed on these tubes.

### 2.5 Molecular Genetics analysis

#### DNA extraction

DNA was extracted from whole blood samples using the saturated salt method. Briefly, 10 mL of RBCs Lysis buffer was added to 2 mL of the collected blood in Falcon tube of 15 mL, that centrifuged at 6000 rpm for 5 minutes the supernatant was discharged, this step was repeated until all RBCS were washed out and clear white pellet formed. 2 mL of white cells lysis buffer was added, 1 mL of Guanidine Hydrochloride, 300 µL Ammonium acetate and 10 µL of Proteinease K (10 mg/ml) were also added. The tube was incubated at 37° C overnight. Pre-chilled 2 mL of chloroform was added to the mixture then vortexed and centrifuged for 5 min at 6000 rpm. The upper layer was added to a new tube and 10 mL of cold absolute ethanol was added, the tube was shaken and incubated for overnight at -20 °C then centrifuged for 10 minutes at 6000 rpm and the supernatant was drained. The pellet was washed with 70% ethanol, and was centrifuged for 10 minutes at 6000 rpm and supernatant was drained. Finally, the pellet which is the DNA was dried from the ethanol then dissolved in 100 µl of distilled water and stored at 4°C for overnight incubation before running the PCR or at -20°C for storage.

### 2.6 Conventional PCR amplification

DNA regions for the *LEF1* exons 2 and 3 were amplified by conventional PCR with an automated thermal cycler (ESCO HEALTHCARE). Primers were obtained from New England Biolabs (UK) Ltd. For exon 2, the primers were forward 5’-TTT TCT TTC TTT TGG GTG TGG -3’ and reverse 5’-AAA TTG CAC CCC TTA TCT GC -3’; and PCR reactions consisted of the initial denaturation for 4 min at 94°C followed by 35 cycles of denaturation at 94°C for 30 sec, annealing at 56°C for 45 sec and extension at 70°C for 55 sec. The terminal elongation was performed at 70°C for 5 min. The amplified PCR product is 140 bp.

While for exon 3, the primers were forward 5’-AAAGGGAAGTCAGTGCATCATT -3’ and reverse 5’-ACAAATCAATTTGCACTTCTGAAC -3’. The PCR reaction mixture was denatured for 3 min at 94°C; then 35 cycles of denaturing at 94°C for 30 sec, annealing of primers at 63°C for 30 sec, extension at 72°C for 45 sec and primer final extension at 72°C for 5 min. The band size of the PCR product is 180 bp.

2% agarose gel was used to visualize the DNA bands and purified PCR products were used for DNA sequencing.

### 2.7 DNA sequencing for exon two and exon three

A twenty PCR products with clear band were sent for sequencing. Sanger sequencing was performed by Macrogen Company (Seoul, Korea). Two chromatograms for each sample, (forward and reverse), were checked for quality and then proceeded for further bioinformatics analysis.

### 2.8 Bioinformatics analysis

#### 2.8.1 Sequence analysis and SNPs detection

The chromatogram sequences were visualized through FinchTV program version 1.4.0 (25). The nucleotide sequences of the studied genes were searched for sequence similarities using nucleotide BLAST NCBI (http://blast.ncbi.nlm.nih.gov/Blast.cgi) (26). Then the nucleotide sequences were translated into amino acid sequences (27). All sequences were subjected to multiple alignment sequences using BioEdit software version 7.2.5 (28); and compared with a reference sequence (LEF1 NCBI Reference Sequence: NG_015798.1, Uniprot protein ID: Q9UJU2) (29) to detect the variants.

#### 2.8.2 Functional analysis of SNPs

##### 2.8.2.1 prediction of the effects of the variants on protein function

We have used three web servers to predict the functional impact of SNPs on the LEF1 protein: 1) Sorting Intolerant From Tolerant (SIFT) server (https://sift.bii.a-star.edu.sg/) which depends on the sequence homology and the degree of conservation for each amino acid in query sequence (30). 2) Polymorphism Phenotyping v2 (PolyPhen-2) server (http://genetics.bwh.harvard.edu/pph2/) which analyzes the multiple sequence alignment and 3D structure of the protein to predict the effect of amino acid substitutions on both protein structure and function (31). 3) Protein Variation Effect Analyzer (PROVEAN) (http://provean.jcvi.org/index.php) which uses BLAST search to find homologous sequences and generates scores for the impact of single and multiple amino acid substitutions, insertions, and deletions on the protein function (32).

##### 2.8.2.2 Prediction of variants on protein stability

I-Mutant 2.0, (http://gpcr2.biocomp.unibo.it/cgi/predictors/I-Mutant3.0/IMutant3.0.cgi), was applied to identify the SNPs effect on the protein stability using the LEF1 sequence. I-Mutant is a neural network-based tool that uses the algorithms of the Support Vector Machine and the ProTherm database (33). It assesses the thermodynamic free energy change upon SNPs in protein sequences.

##### 2.8.2.3 Prediction of variants pathogenicity

In this study, three web servers have been used to evaluate the pathogenicity score of the variants to touch upon: 1) MutationTaster application (https://www.mutationtaster.org/) which integrates information from different biomedical databases and uses established analysis tools (34). Analyses include loss of protein features, splice-site changes, changes that might affect the amount of mRNA and evolutionary conservation. MutationTaster employs a Bayes classifier to eventually evaluate the disease potential of DNA sequence alterations. 2) PMUT which uses a neural network-based classifier trained by a manually curated dataset extracted from SwissProt (35) to predict the pathological nature of a given single amino acid variant, and also hot-spot positions on protein sequences (36). 3) MutPred2 software which classifys amino acid substitutions as pathogenic or benign in human depending on their impact on over 50 different protein properties and, thus, enables the inference of molecular mechanisms of pathogenicity (37).

#### 2.8.3 Splicing analyses of exonic three mutations of *LEF1*

Splicing analyses were performed with different bioinformatics web-based tools to identify mutations that have potential effect in pre-mRNA processing. Splice sites have been predicted by neural network (NNSplice) (https://www.fruitfly.org/seq_tools/splice.html) and NetGene2 (https://services.healthtech.dtu.dk/service.php?NetGene2-2.42) (38-40). In addition, to investigate the presence of potential splicing regulatory sequences, i.e. ESEs and ESSs, and identifiy the potential effect of mutations on splicing regulatory motifs, we used ESEfinder 3.0 (http://rulai.cshl.edu/tools/ESE2/index.html) and FAS-ESS (http://hollywood.mit.edu/fas-ess/) (41, 42). Also, MutationTaster and CRYP-SKIP servers (https://cryp-skip.img.cas.cz/) were used to identify exonic mutations that could possibly affect pre-mRNA splicing leading to disease and distinguishes the two aberrant splicing outcomes from DNA sequences, respectively (34, 43).

#### 2.8.4 Homology Modeling

A predicted 3D structure of the *LEF1* protein sequence (ID: Q9UJU2) was obtained from RaptorX Property, a web server, (http://raptorx2.uchicago.edu/StructurePropertyPred/predict/) (44). Then UCSF Chimera (version 1.8), which is available from the chimera web site (http://www.cgl.ucsf.edu/cimera) (45), was used to visualize the 3D structure.

## 3. Result

### 3.1 Characteristics of the study population

A total of 128 chronic lymphocytic leukemia patients, 104 (81.3%) males and 24 (18.7%) females, with a mean age of 63.8 years were enrolled in this study. The clinic-pathological characteristics of patients are summarized in Table 1.

**Table 1.**
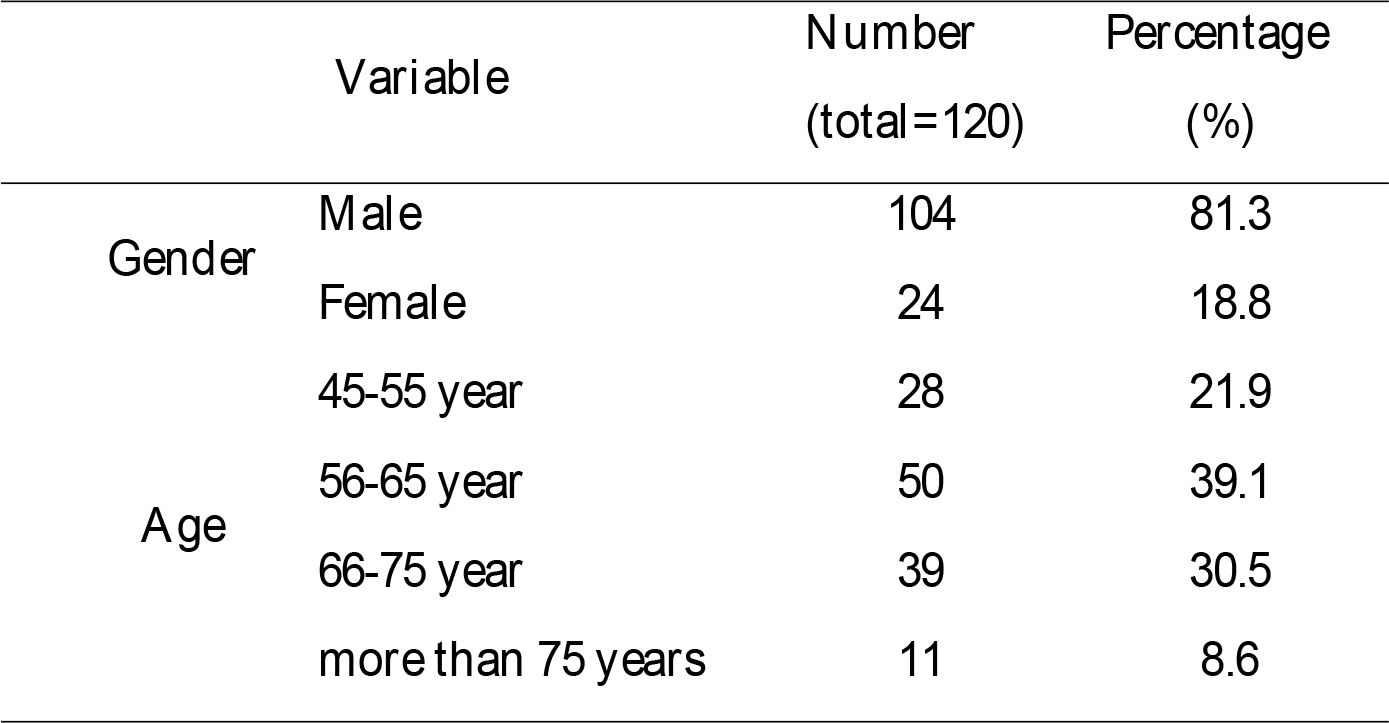
The clinic-pathological characteristics of patients

| Variable |  | Number<br>(total=120) | Percentage<br>(%) |
| --- | --- | --- | --- |
| Gender | Male | 104 | 81.3 |
|  | Female | 24 | 18.8 |
| Age | 45-55 year | 28 | 21.9 |
|  | 56-65 year | 50 | 39.1 |
|  | 66-75 year | 39 | 30.5 |
|  | more than 75 years | 11 | 8.6 |

### 3.2 Complete blood count (CBC) result

Complete blood count (CBC) measured with a hematology analyzer which revealed 89.4 X103/L. Leukocyte count (WBC), Red blood cell count (RBCs), Platelets count (Plts) and hemoglobin (Hb) were reported in Table 2.

**Table 2.** Measures of Leukocyte count, red blood cell count, Platelets count and hemoglobin

|  | Total<br>number | Range | Mean | Std. Deviation |
| --- | --- | --- | --- | --- |
| WBCs | 128 | 89.4 x 10<sup>3</sup> | 96713.3594 | 113247.00011 |
| Hb | 128 | 10.80 | 10.7602 | 2.26181 |
| Plts | 128 | 918.00 | 168.1961 | 118.33119 |
| RBCs | 128 | 5.20 | 3.8414 | .98385 |

WBCs: Leukocyte count, RBCs: Red blood cell count, Plts: Platelets count and Hb: Hemoglobin.

### 3.3 Flow cytometry result

All of the 128 patient samples were analyzed using flow cytometer technique to evaluate their CD5 and CD19 markers which are the main markers for CLL cells, firstly; the samples examined against forward and side scattered light to visualize the size and granulation/vaculation of the cells which this step showed that the cells are located in lymphocytes area, secondly; addition of CD45 and examine against side scatter only to predict the type of lymphocytic cells and our results were posited in the area of small lymphocytes (mature lymphocytes), thirdly which is the last step; addition of CD5 and CD19 to the samples; the results were high peak in the strong positive area of CD5 when added alone to the samples, also high peak in the strong positive area of CD19 when added alone to the samples and most of the cells occurred in double positive area when visualized against CD5&CD19 together. The graphs are illustrated in Figure 1.

**Figure 1.**
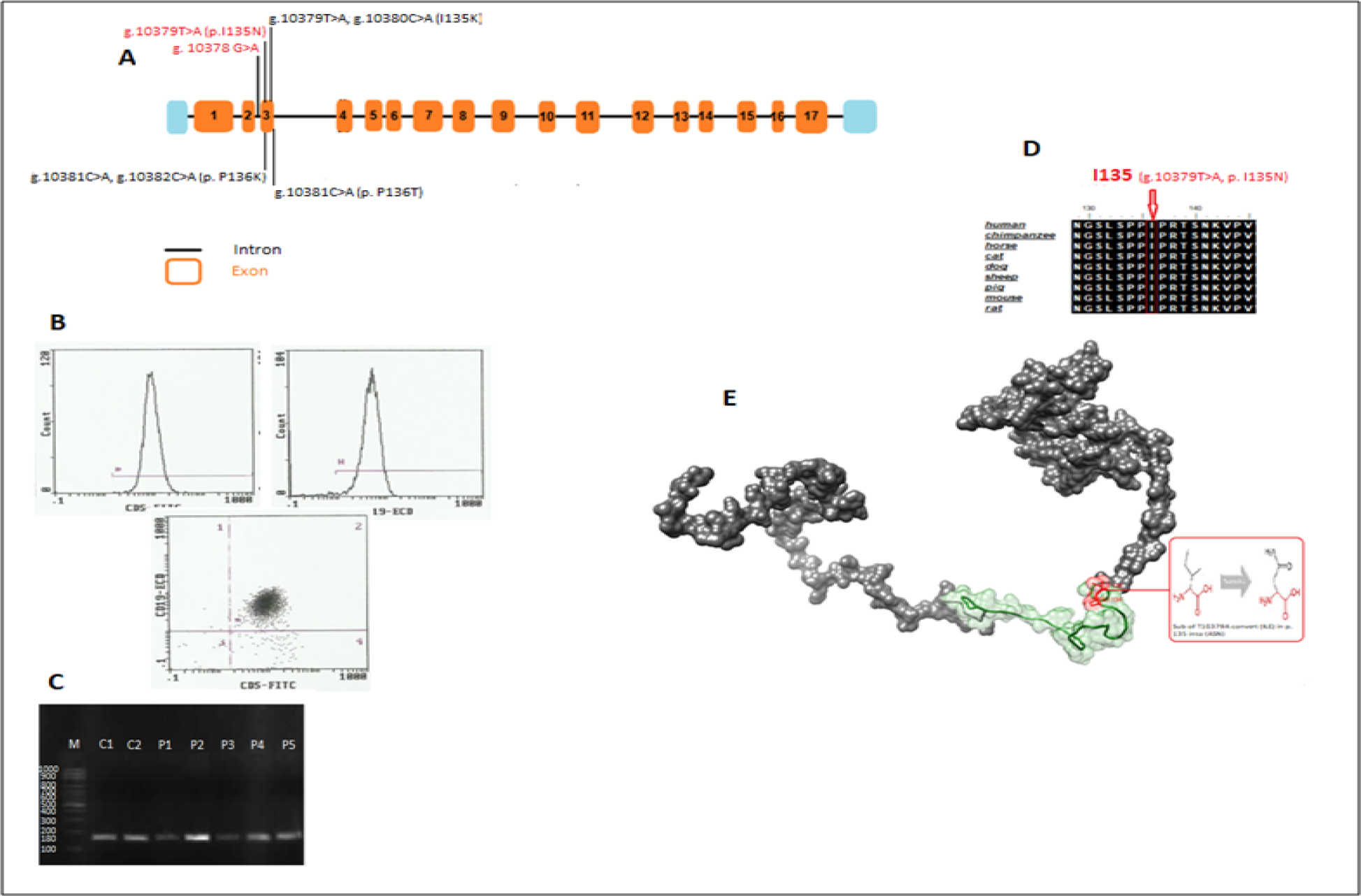
CD5&CD19 and mutational analysis of *LEF1* gene in Sudanese CLL patients. (A) Scheme of *LEF1* gene showing the localization of all identified mutations along its coding sequence. Exons are numbered. Common and Novel mutations are highlighted in red. (B) Showing of peaks that in a highly strong area of CD5&CD19 and the double positivity results for both markers. (C) Displaying of Conventional PCR results visualized by gel electrophoresis that showing the bands of positive controls and the patient samples against specified marker (ladder) to determine the size of the sample DNA sequence which is determined to be 180 bp. (D) Alignments of nine vertebrate amino acid sequences of *LEF1* demonstrating that the residues predicted to be mutated in our Proband (indicated by the red arrow) are evolutionarily conserved across species. (E) Three-dimensional structural modeling of *LEF1* gene with a showing of LEF1 native amino acid (Ile) that affected by the substitution of T10379A and convert it into Asn in position 135 (using the Chimera 1.8 program).

### 3.4 Functional analysis of nucleotide variations in *LEF1*

When we screened mutations of *LEF1* exon two and exon three, the following sequence variants were observed in comparing with the reference sequence (NG_015798.1). In exon two, we did not find any mutation but we detect one (NG_015798.1:g.10178G>A) in intron two in 12 patients. While in exon 3, we found four mutations: NG_015798.1:g.10377C>A, g.10379T>A, g.10380C>A, and g.10381C>A. However, the presence of these mutations differs from one patient to another. All of 20 tested patients carry these mutation g.10377C>A and g.10379T>A. The majority of them, 8 patients and 6 patients, have g.10380C>A andg.10381C>A, respectively. The only one patient carries all of the observed mutations. Accordingly, the patterns of resulted amino acid are different. See Figure 2 and Table 3 for more explanation. The function analyses of the resulted amino acid variants, as predicted by several in silico tools, are presented in Table 3 and Figure 1.

**Table 3.** Nucleotide variations of exon three of *LEF1* observed in B-CLL Sudanese patiens

| Nucleotide changes<sup>a</sup> | HGVS coding | HGVS protein level<sup>b</sup> | rs ID |
| --- | --- | --- | --- |
| g.10377C>A | NM_016269.4:c.402C>A | p.Pro134Pro | - |
| g.10379T>A | NM_016269.4:c.404T>A | p.Ile135Asp* | - |
| g.10379T>A, g.10380C>A | NM_016269.4:c.404_405TC>AA | p.Ile135Lys* | - |
| g.10381C>A | NM_016269.4:c.406C>A | p.Pro136Thr | rs1048060629 |

<sup>a</sup> Position is determined according to Ref sequence (NG\_015798.1)
<sup>b</sup> Position is determined according to Ref sequence (NP\_057353.1)
\* Novel mutations detected in this study

**Table 4.**
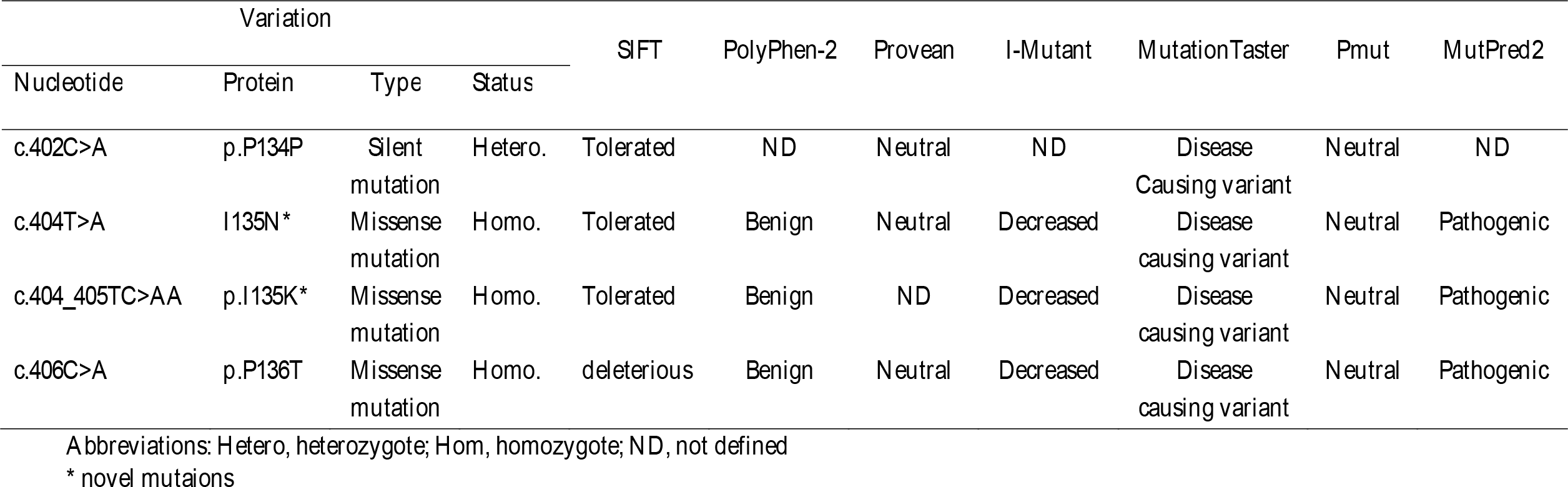
Several online servers that used to functionally analyzed detected variants in exon three of LEF1 in B-CLL Sudanese patients. The annotation was applied according to the Homo sapiens genome assembly GRCh37 (hg 19).

| Variation |  |  |  | SIFT | PolyPhen-2 | Provean | I-Mutant | MutationTaster | Pmut | MutPred2 |
| --- | --- | --- | --- | --- | --- | --- | --- | --- | --- | --- |
| Nucleotide | Protein | Type | Status |  |  |  |  |  |  |  |
| c.402C>A | p.P134P | Silent mutation | Hetero. | Tolerated | ND | Neutral | ND | Disease Causing variant | Neutral | ND |
| c.404T>A | I135N* | Missense mutation | Homo. | Tolerated | Benign | Neutral | Decreased | Disease causing variant | Neutral | Pathogenic |
| c.404_405TC>AA | p.I135K* | Missense mutation | Homo. | Tolerated | Benign | ND | Decreased | Disease causing variant | Neutral | Pathogenic |
| c.406C>A | p.P136T | Missense mutation | Homo. | deleterious | Benign | Neutral | Decreased | Disease causing variant | Neutral | Pathogenic |

Abbreviations: Hetero, heterozygote; Hom, homozygote; ND, not defined
\* novel mutaions

**Figure 2.**
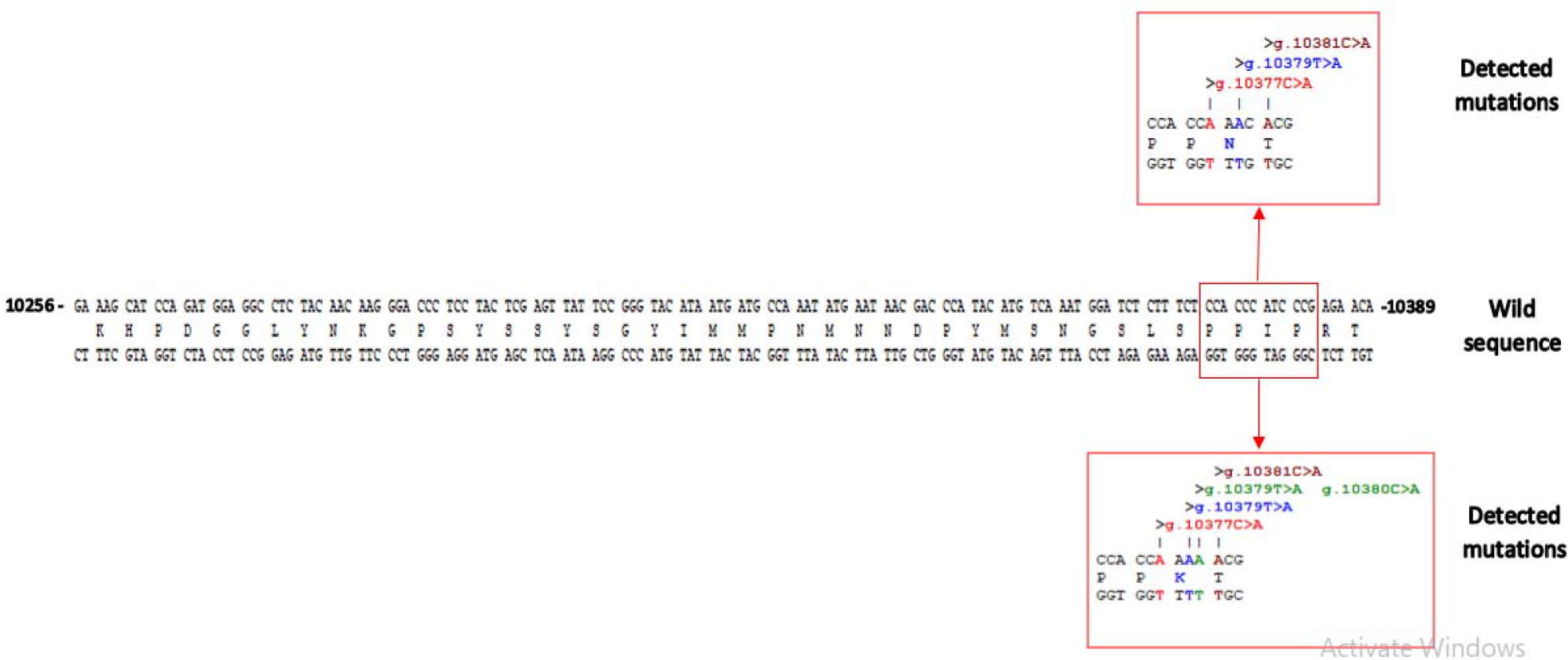
The pattern of genetic alterations in exon three of the *LEF1* gene and their corresponding amino acid in our studied patients. The position is determined according to the reference sequence (NG_015798). The figure was taken from Serial Cloner (v.2.6.1).

### 3.5 The splicing analyses of exonic three mutations

NNSplice software predicted two 5′ splice donor sites, at position 10314 and 10390 with 0.57 and 0.94 scores, respectively, and one 3’ splice acceptor at position 10255 with 0.91 prediction score. While NetGene2 tool predicted one 5′ splice donor sites at position 10390 with 0.83 score, and one 3’ splice acceptor at position 10255 with 0.56 prediction score (position is determined according to reference sequence NG_015798.1). However, ESEfinder predicted 20 ESE consensus motifs that are recognized by one of four different SR proteins (SF2/ASF, SC35, SRp40 and SRp55). Also, it predicted that the four mutations, c.402C>A, c.404T>A, c.404_405TC>AA, and c.406C>A, could involve in the alteration of an ESE sites. Moreover, CRYP-SKIP servers showed that the presence of these mutations could lead to exon skipping (probability of cryptic splice site activation (*P*CR-E) is equal to 0.41. It takes values between zero and one, with higher values (<0.5) speaking in favor of cryptic splice-site activation and lower values (>0.5) in favor of exon skipping. The results of splicing analyses of the four mutations, that located in *LEF1* exon three, are illustrated in Table 5.

**Table 5.** Splicing analysis of four mutations located in LEF1 exon three using web- based in silico tools

| Mutation | Position<sup>a</sup> | FAS-ESS | ESE Finder | Mutation Taster |  | CRYP-SKIP |  |
| --- | --- | --- | --- | --- | --- | --- | --- |
|  |  | Gained ESS<sup>b</sup> | Disrupted ESE<sup>b</sup> | prediction | effect | Splice Effect | *P*<sub>CR-E</sub> |
| p.P134P | 13 | 0 | 2 | Disease causing | Donor gained | EXSK | 0.44 |
| I135N | 11 | 0 | 1 | Disease causing | Donor increased | CR-E | 0.54 |
| p.I135K | 10 | 0 | 2 | Disease causing | Acc increased<br>Donor increased | EXSK | 0.47 |
| p.P136T | 9 | 0 | 1 | Disease causing | Acc marginally increased | CR-E | 0.52 |

Abbreviations: ESE, Exonic Splicing Enhancer; ESS, Exonic Splicing Silencer; FAS-ESS, fluorescence-activated screen for ESS; Acc, acceptor; EXSK, exon skipping; CR-E, cryptic splice-site activation; *P*<sub>CR-E</sub>, probability of cryptic splice site activation.
<sup>a</sup> Position of variant relative to the nearest splice site, 10390.
<sup>b</sup> Values 0, 1, 2 and 3 indicate number of splicing regulatory elements gained or disrupted

## 4. Discussion

Chronic lymphocytic leukemia (CLL) is the most common hematologic malignancy with a unique disease trajectory (47). It is now recognized as a heterogeneous disease with a variety of clinical outcomes. Despite the effectiveness of chemoimmunotherapy (CIT), most CLL patients after starting the treatment will relapse at some point within the first five years (48). However, many factors are responsible for CLL relapse and patients’ responses to the treatment (49). Mutations in the essential genes involved in lymphocyte development and involved in lymphocyte development, such as the *LEF1* gene, are among the high-risk factors for CLL relapse and treatment response (50-52). In this study, we performed mutational analysis for *LEF1* exons two and three, the hotspot regions in leukemia (24), in Sudanese CLL-patients, and we identified four mutations in exon three, two of them were not reported previously. Interestingly, splicing analysis predicted that these mutations could lead to splicing defects in *LEF1* pre-mRNA due to their potential effects on splicing regulatory elements (i.e. ESE).

The literature reveals that the exonic sequences are involved in the regulation of pre-mRNA splicing in addition to their protein-coding potential (53-55). Because exons contain cis-regulatory elements, such as exonic splicing enhancers (ESEs) and exonic splicing silencers (ESSs) which promote or inhibit the recognition of the neighboring splice sites, respectively (53). In addition, in many circumstances in which pre-mRNA splicing is affected by an exonic mutations, it is known that ESE elements are commonly present in exons to assess in identifying weak splice sites (53, 56). In this study, we found that the four mutations lead to disrupt the splicing regulatory elements, ESE (Table 5). Also, the mutations were detected in patients with high WBC, range from 18.2×10^9^/L to 900×10^9^/L) at pretreatment. This finding is in agreement with previous studies which reported *LEF1* mutations in *LEF1* gene and related them to high risk events in chronic lymphocytic leukemia/small lymphocytic lymphoma (CLL) (24, 57). However, Sonja and his colleagues have confirmed, in their genome wide association (AGWs) study, the association between *LEF1* gene polymorphisms and an increased risk for CLL (21). Moreover, the two mutations Pro134Pro and Ile135Asn (novel mutation) were detected in all patient and could be used as diagnostic and/prognostic markers for CLL. Regarding the LEF1 protein properties, the conversion of isoleucine into an asparagine at position 135 makes changes that related to amino acid properties. The mutant is bigger in size and less or totally lost of hydrophobic interaction than the wild type (58). Although substitution of Pro134Pro performs no change in proline amino acid at position 134, this synonymous variant is also reported in International Cancer Genome Consortium (ICGC) somatic mutation databases in association with hepatocellular carcinoma (59, 60).

A positive CD5 & CD19 is one of the most diagnostic markers for the presence of chronic lymphocytic leukemia. We detected the B cells CD5 and CD19 markers by flow cytometry technique in our patient’s samples and they were strongly positive and located in the highly positive area. In a study conducted in 2016, Wei Wu and his colleagues evaluated the CD markers in different samples (core blood, bone marrow, peripheral blood …etc) in CLL patients and they also found a positive CD5&CD19 for patients’ peripheral blood test (61). Moreover, CD19+ is considered, by the Matutes score, as one of the well-defined immunophenotype markers for CLL while CD5 can be occasionally co-expressed in other lymphomas, such as lymphoplasmacytic lymphoma (LPL) and follicular lymphoma (FL) (62-65). The present study was found a positive correlation between CD5 and CD19 expression; and *LEF1* mutations in B-CLL Sudanese patients. *LEF1* gene expression has been identified to be expressed by lymphocyte B cell precursors (57). Albert *et al*. reported that the *LEF1* is over expressed in CD5+/CD19+ B cells from CLL patients compared to those from healthy donors (19).

The limitations of the study include the relatively small number of the enrolled patients, and depending on the bioinformatic tools to assess the possible effect of the identified changes. We detected the two mutations Pro134Pro and Ile135Asn (novel mutation) in all enrolled CLL patients and they could be used as diagnostic and/prognostic markers for CLL. Therefore, further in vitro and in vivo functional studies with large sample size are required to verify the splicing effect of the detected mutations in *LEF1* pre-mRNA.

**In conclusion**, in this study, we found a positive correlation between CD5 and CD19 expression; and *LEF1* mutations in CLL Sudanese patients. Four mutations in exon three, two of them were not reported previously were identified. Interestingly, splicing analysis predicted that these mutations could lead to splicing defects in LEF1 pre-mRNA due to their potential effects on splicing regulatory elements (i.e. ESE). Also, we proposed that the *LEF1* could be used as useful marker in the diagnosis of CLL; and there is an association between *LEF1* gene mutations and an increased risk for CLL. However, further studies with large sample size are recommended to confirm our findings.

## Data Availability

All data produced in the present work are contained in the manuscript

## Ethics statement

Ethical clearance was taken from the Ethical Review Board of the National Ribat University. Informed consent was obtained from each patient. The confidentiality of patients was kept as no names were recorded. The participants were provided with information about the study and any risk that may arise especially when the collection technique was applied.

## Acknowledgments

We thank the staff of Radiation and Isotopes Centre of Khartoum (RICK) for their kindly contribution and all participants in the study.

## Data availability

All data produced in the present work are contained in the manuscript.

